# Brazilian model estimation for SARS-CoV-2 peak contagion (BMESPC)

**DOI:** 10.1101/2021.01.02.20248940

**Authors:** Guilherme Asai, André Kuroiva, Manuella Lucca Terra

## Abstract

With newer data for SARS-CoV-2 and entering the second wave of contagion required the improvement of the forecasting model, structuring its model to forecast the peak of the first and second contagion wave in Brazil. The Brazilian model estimation for SARS-CoV-2 peak contagion (BMESPC) was structured, capable of estimating the peak of contagion for SARS-CoV-2 in the first and second waves, as the main objective of this work. Using the BMESPC model, it was possible to estimate, with a certain reliability degree, the peak of contagion for the first and second waves in Brazil, with one day difference from the real to the forecast. While at the state level, the calculated confidence interval proved to be more accurate. In this way, it is possible to use BMESPC to forecast the peak of contagion for several regions, provided that the necessary structure and calibration are respected.

## Introduction

Approximately one year after the first people were infected by a new virus in Wuhan County in China, the world started to face a growing wave of contamination that, hourly fluctuates negatively, hourly positively. Despite all the measures, health, and policies, taken, the outbreak is far from controlled. In many places, there is a re-opening of the economy which can cause an increase in the number of cases of the disease.

In August, September, and October, some countries chose to relax the quarantine. In the same period, the number of confirmed cases, which was in decline, rose again. The world is sensing an increase of SARS-CoV-2 (Covid-19) contamination with several countries such as France, Spain, and the United Kingdom that are reporting an increasing number of new Covid-19 cases. This increase in cases is due to the second wave of contagion as point some studies like Xu and Li (2020), Cacciapaglia, Cot, and Sannino (2020), and Vogel (2020).

In some countries, the increase in new cases coincides with economic opening with the opening of local commerce and leisure areas. The initial decrease in the number of new cases of Covid-19 encourages countries to eased physical isolation because some locations observed a drop in the number of new cases accompanied by the release of ICU beds in hospitals. This caused a false sense that the pandemic was under control and leads to easy physical isolation at this moment.

After a period of decrease in number of confirmed cases, at the beginning of the second semester, some countries like the USA, India, Argentina, and Russia report an increase in Covid-19 cases for a few days during July 2020 and a more significant increase during September. France and Spain, countries where they had a major outbreak in Europe in the first quarter of 2020, recorded consecutive increases in cases throughout September and October.

With the increase in the number of new cases and the concern about a new wave of contagion, it has led the government of some countries to adopt restrictive measures of isolation and setback in opening the economy. In some parts of Spain and Germany, the lockdown returned and in the UK there was a restriction on the opening of pubs, for example. However, restrictive measures took place, especially in European countries.

Figure 1 illustrates the new daily case growth of Covid-19.

**Figure 1.**
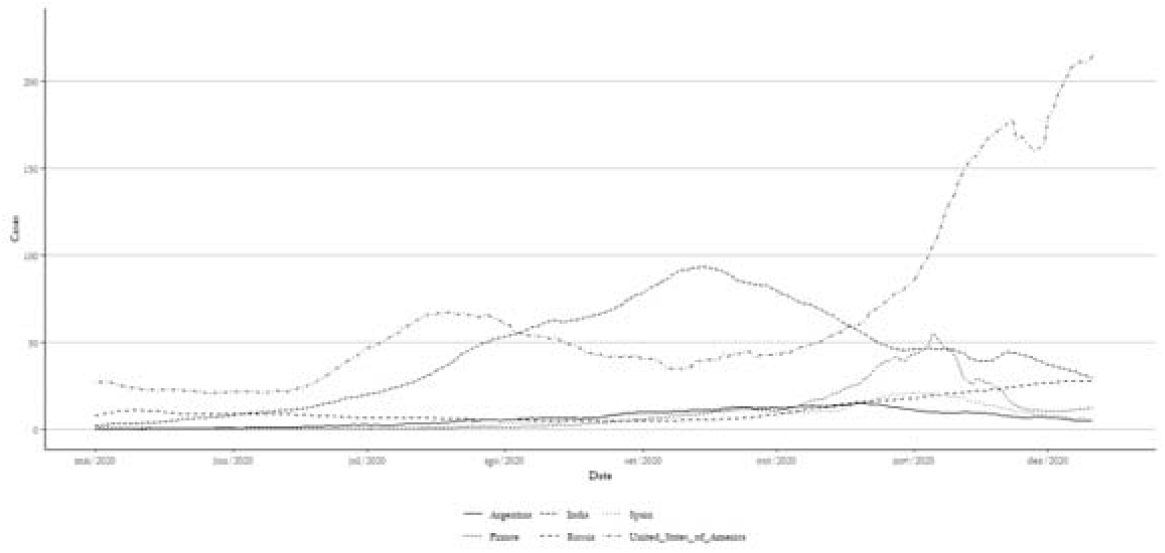
A 7-day moving average of SARS-CoV-2 new cases in selected countries between May and early mid-December (cases x 1,000). Source: Our World in Data – Coronavirus (COVID-19) Cases, 2020.

When looking at Figure 1, the USA shows a significant increase in the number of cases between November and December. If you look at the USA in isolation, two waves of contagion are visible, the first May to September and the second beginning in October.

By this time, Brazil continues at a pace of growth in the number of new cases after a three-times plateau stabilization between May 5^th^ to June 9^th^, June 19^th^ to July 9^th^ and, August 27^th^ to September 10^th^. After each time of flattening of the contagion curve, loosening measures of the quarantine, motivated the increase in the number and peaks. Over time, a decrease in the number of new cases was observed from September to October and all Brazilian states loosened the physical isolation measures. Nevertheless, it is noted that, after the quarantine flexibility dates, with the opening of shops, bars, restaurants, and leisure areas, there was an increase in the number of cases and Brazil reach the Covid-19’s second wage of contagion.

Figure 2 indicates a moving average for the new daily case growth of Covid-19 in Brazil.

**Figure 2.**
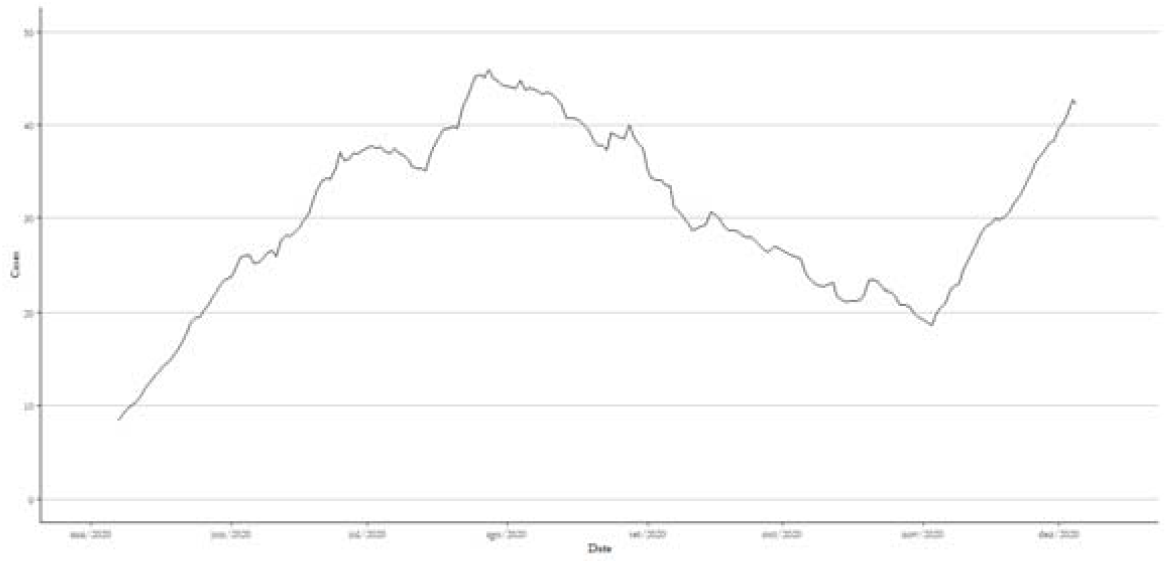
A 14-day rolling average for SARS-CoV-2 contagion in Brazil (cases x 1,000). Source: author’s elaboration. Data: Our World in Data – Coronavirus (COVID-19) Cases, 2020.

As in the USA, it is possible to distinguish two contagion waves in Brazil. the first between May to mid-October and the second beginning in November. The turning point of the curve in November symbolizes the flexibility of the quarantine in several states and municipalities, whose rate of physical isolation has returned to pre-pandemic levels.

Also, the holidays that followed in the months of August, September, and October, combined with the good wheatear, instigated many people to go to the beach in a pandemic time. After these periods, when there was a greater agglomeration of people, the number of cases in Brazil slightly increased.

According to Dandekar and Barbastathis (2020), Palomino and Budini (2020) and Hornstein (2020) the relaxation of the quarantine has to be different levels and step by step. In each location the quarantine must be treated differently, the main point being the speed and way of detecting and isolating the individual with the disease so that relaxation occurs. To maintain control of contagion, it is necessary to establish scenarios that allow the monitoring of the progress of the disease and for the economy to open up. However, it is difficult for public policies to comply with the recommendations imposed by health agencies since there is external pressure for the economy to return to work quickly. At the peak of the pandemic, Piguillem and Shi (2020) indicate that the ideal isolation would be greater than 70%, however, in many locations, the index was between 20 to 50%. Zhao and Chen (2020) add that measures such as travel restraint and isolation of suspected cases are indicated to contain the pandemic’s progress.

While these measures point to control the SARS-CoV-2, another is important for easing quarantine. To Colbourn (2020) to lockdown, measures are important to countries to incorporate testing, contact tracing, and localized quarantine of suspected cases. Doing these steps is possible for planning a relaxing social distance.

With this discussion about flexibilities or continuing quarantine, even with an increasing number of newer cases of Covid-19 plus the possibility of new contagion waves, it becomes important to carry out a forecast of the peak of contamination. This can help public policies in planning the relaxation of quarantine.

### Objective

Structure a forecasting model capable of predicting the SARS-CoV-2 peak of contagion for the first and second waves in Brazil and that can be replicated for any region.

### Brazil’s SARS-CoV-2 disease background

The Covid-19 outbreak started in Brazil on February 26^th^, when the city of Sao Paulo registered the first case. After ten days, on March 5^th^, there was the first case of local transmission, that is, the first case where a Brazil’s resident contracted the virus in the national territory, also in the city of Sao Paulo. In September, the state of Sao Paulo was the first state in the world to reach the mark of 1 million contaminated by Covid-19.

Nevertheless, the Sao Paulo State is not the only one in the country that concerns national and international health authorities. Other states like Rio de Janeiro, Bahia, Amazonas, and Minas Gerais have had many confirmed cases. More recently, the state of Mato Grosso and Mato Grosso do Sul showed high indications, and especially in the indigenous population.

Numerous studies list the determining factors for the spread of the virus, these determinants are influenced by economic to environmental issues. Since it is a respiratory disease, the factor of climate and geographic region interferes with the speed of dissemination as suggested by Shi *et al* (2020) and Rhodes *et al*. (2020). Both studies suggested that the climate affects the virus spread, if the local has more humidity and tropical temperature more cases were observed. Prata *et al*. (2020) indicate that there is a negative linear relationship between temperatures and daily cumulative confirmed cases, which means that lower temperature, decreases the of Covid-19 confirmed cases.

Socioeconomics factors are also a determining factor to spread Covid-19. Racial disparity, population density, economic development, individual economic conditions, access to hospitals and, the number of UCIs are risk factors to contamination in which people in a given region are exposed to contagion the lower their economic and social status. (Mclaren 2020, Patel *et al*. 2020, Chowkwanyun and Reed 2020).

By combining these factors, Brazil tends to be a country with characteristics for the rapid spread of the virus, it is no wonder that in a short time it became the third country with the largest number of confirmed cases, right after the USA and India.

For having continental proportions and also for having different socio-economic and climatic characteristics, Brazil has different behaviors among its macro-regions. This behavior affects the speed of propagation of the virus and also the number of people infected. According to Prata *et al*. (2020), the climate is a factor that interferes in the spread of the virus, and in milder climates, the speed of contagion decreases. This fact is observed in the southern region of Brazil, where the level of contagion is lower than the rest of the regions.

Figure 3 shows the moving average of new daily cases for the Brazilian five macro-regions.

**Figure 3.**
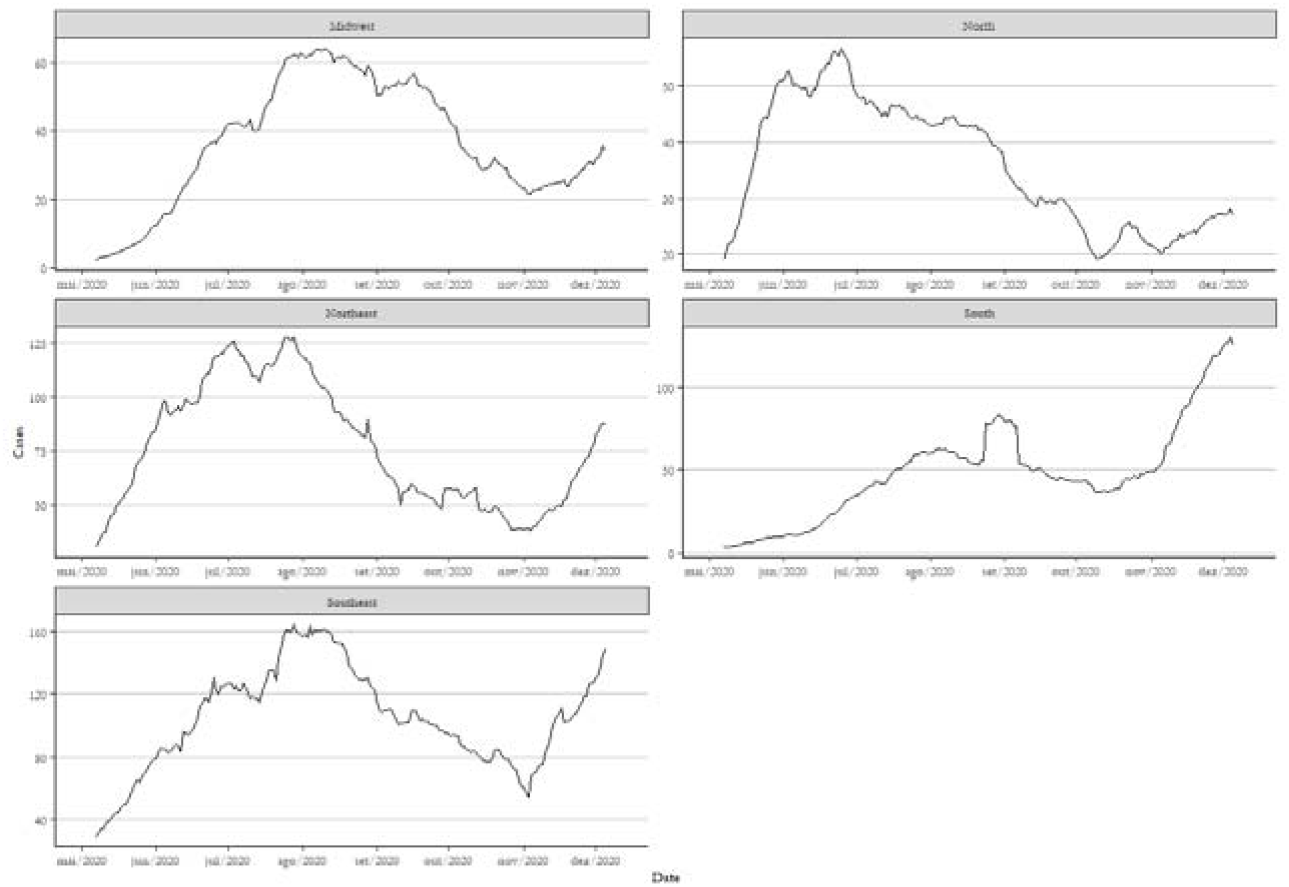
A 14-day rolling average for SARS-CoV-2 contagion for Brazilian macro-region (cases x 1,000). Source: author’s elaboration. Data: Ministério da Saúde, 2020.

This overview shown in Figure 3 indicates the disparity of Covid-19’s cases in Brazil. The southeastern region has become the epicenter of the pandemic in brazil due to its high population density and also the high traffic of people between the main cities of the country (Sao Paulo, Rio de Janeiro, and Belo Horizonte). Even with the restriction of travel (plane and bus) to these cities, the regions continued to receive a large flow of people that contributed to the spread of the virus and the increased contagion between the states in the Southeast.

In turn, the northern region has the least number of cases. This is due to the low population density present in the states, that is, the agglomeration of people is not as much as in the southeast. However, at the beginning of the sanctuary crisis caused by Covid-19, the state of Amazonas, in particular, its capital Manaus, suffered from the collapse of the health system and the funerary system, whose large volume of cases and deaths caused lockdown measures. The northeastern region, the second in number of cases, was another that adopts lockdown measures due to a large number of cases and deaths.

To contain the outbreak of the Covid-19and prevent the health system from collapsing with the high UCI occupancy rate, the government, at the state level, structured physical distance measures (social distance), closing areas of crowding of people like pubs, restaurants, parks, shopping malls and restricting streets commerce.

After this initial concern, government officials have relaxed the quarantine, with the opening of the economy and the liberation of the agglomeration area. Coincidentally, with these measures, cases of contagion, which remained on a plateau, began to increase. It is inaccurate to say that the easing of the quarantine with the economic opening is the main factor for the increase in cases, but there are indications for this.

Several studies like Painter and Qiu (2020), Block *et al* (2020), and Thunström *et al* (2020) indicates that physical isolation (social distance) measures are efficient to flatten the contagion curve and is more effective when the quarantine is respected and prolonged, even if at the expense of economic losses. Flattening the contagion curve and decreasing the spread of the virus should be a priority to contain the progress of the disease.

Thereby, the effects of quarantine and the pandemic are felt in the economy as a whole, both in the short and long term. Studies such as those in Atkeson (2020), Baker *et al*. (2020), and McKibbin and Fernando (2020) indicates that there is a great tendency to recession, with an increase in the unemployment rate, closing deals, decreasing in GDP, less domestic and international supply chain, in addition to increased inflation.

Even for countries with a strong economy, such as the USA, the economic impacts would be great. Baker estimates that the economic uncertainty stemming from the Covid-19 crisis would hover around the 20% drop in GDP for the last four months of the year.

The Brazilian economy also suffers from the impacts of the pandemic. According to Asai and Correa (2020), Brazil could go through a systemic crisis that would impact social welfare, drop in GDP, and decrease wages. The authors estimate that the drop in GDP would be in the order of 700 billion, which would bring the country back to 2007 GDP levels.

Pressured by these factors plus municipal elections, whose measures of social isolation have become unpopular, Brazilian politicians opted for the reopening of the economy. Figure 4 illustrates the actual situation in Brazil after the economic reopening per state.

**Figure 4.**
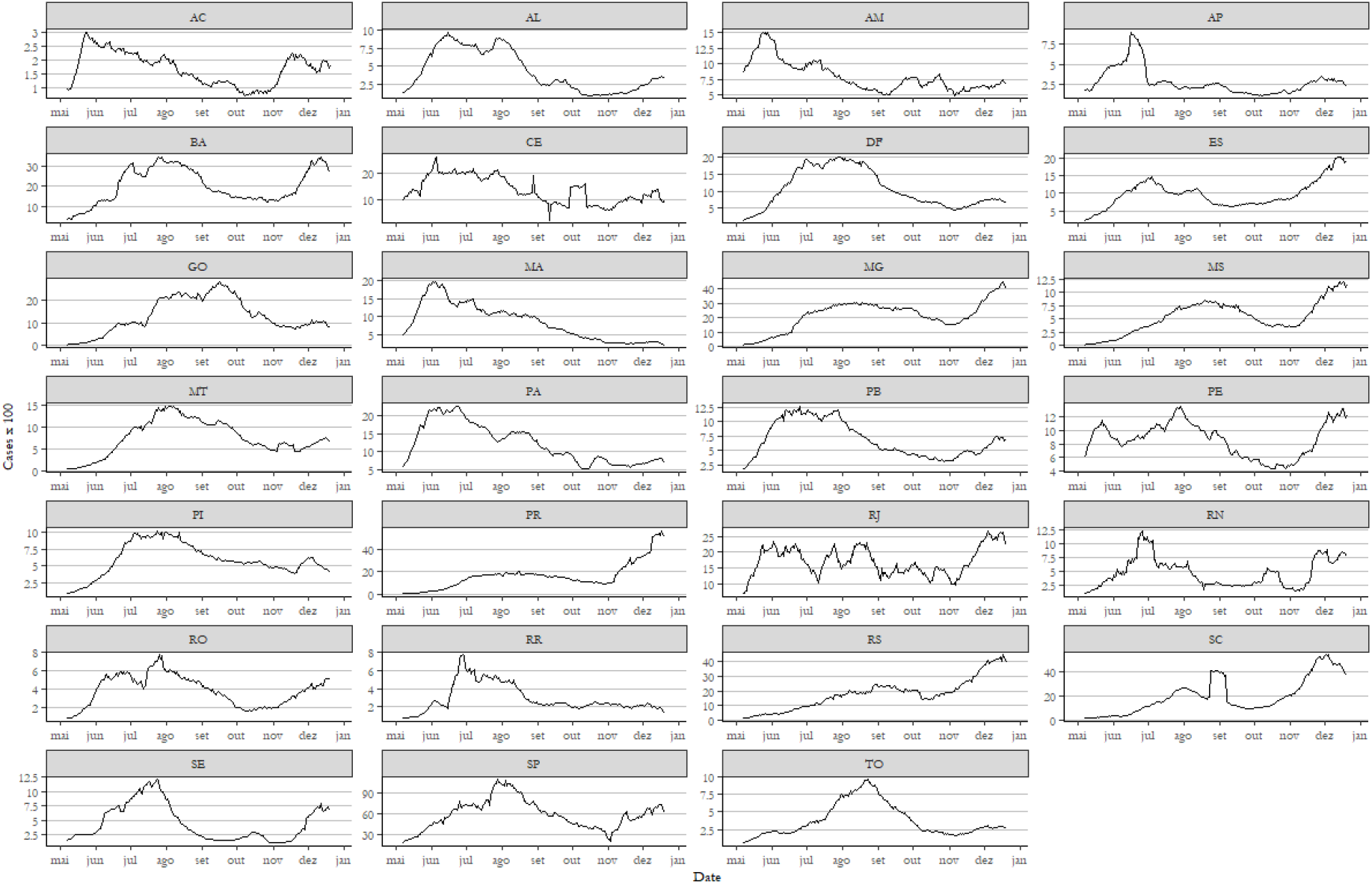
A 14-day rolling average for SARS-CoV-2 contagion in Brazil per state in 2020 (cases x 100). Source: author’s elaboration. Data: Ministério da Saúde, 2020.

Note that there is still an increase in the number of cases in some states, while others reach a plateau or decline. Figure 4 also indicates that there is no standard for spreading the virus, and it is necessary for each state to adopt measures that suit them at the moment and not a single measure.

Therefore, it is important to estimate when the peak of contagion will occur to provide guidelines and a scientific basis for formulating public policies and making decisions about the economic reopening and the flexibility of the economy.

### Analytical approach

The Brazilian model estimation for SARS-CoV-2 peak contagion (BMESPC) is held on Bayesian statistics based on Monte Carlo Method (MCM) that will be used to forecast the contamination peak in Brazil by SARS-CoV-2 (Covid-19), through a Markov Monte Carlo algorithm. Combined with MCM, a Bootstrap Method (BM) will be performed to cross-validation the results. Both methods are considering a 95% confidence interval.

A Markov Monte Carlo algorithm is a usual method to predict or forecast under uncertainty, which attempts to simulate draws from some complex distribution with previous sample values to randomly generate the next sample value, based on the most recent value during the complex distribution of probabilities.

The base MCM model was developed in the field of physics, using random number generation to solve integrals problems. Equation 1 indicates the MCM based integration model.

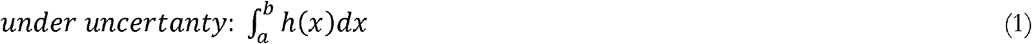

This base integration can be decomposed into a probability function and a density function. In statistical terms, the integration can be expressed as an expectation of a distribution function overdensity as expressed in Equations 2 and 3.

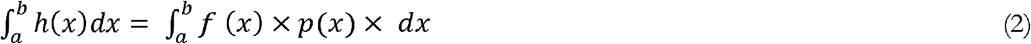

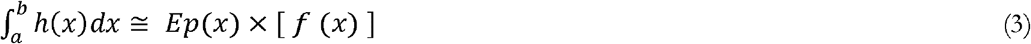

Due to a large number of random variables in *x* the density of *p*(*x*) can be estimated following Equation 4.

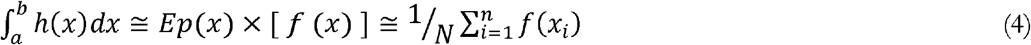

Those four Equations are the base principles of the MCM model which is following in this study. Meanwhile, the complete MCM model used can be found in the work of Geyer (1992), Kalos and Whitlock (2009), and Rubinstein and Kroese (2016).

To randomize the *x* variables, a base distribution to input in MCM was estimated in terms of normal distribution likely *C*∼*N*(*σ*^2^). So, it is possible to create the probability distribution function, as well as the density function needed.

To establish *μ* and *σ* some comparable data was collected. This data refers to the number of days that the country reaches the peak of contamination by Covid-19. Then, it was selecting some countries taking the following criteria: (i) age pyramid similar to Brazil, that is, with the majority of the young population; (ii) more than 20,000 cases; and (iii) data reliability. If the county fulfills all three criteria, it was considered a comparable country. Also, some major countries were added as comparable countries due to their representativeness in the Covid-19 pandemic.

The comparable countries using to previous sample values in the MCM model are United States, United Kingdom, United Arab Emirates, France, Belgium, Germany, Brazil, Canada, South Africa, India, China, Russia, Chile, Sweden, Indonesia, Israel, Peru, Thailand, Bangladesh, Poland, Morocco, Iran, and Ecuador.

To measure the number of days that the country reaches the peak of contamination, it is important to distinguish the waves of contagion, because some of the comparable countries already reach two waves, and others do not. Chile, Ecuador, India, Iraq, Morocco, Poland, and Thailand don’t reach the second wave yet.

There is no particular formula found and the medical literature to identify the contagion waves. According to Johns Hopkins Medicine (2020), the diseases can be seasonal and present waves of contamination that can be identified when the number of infections rises and then decline, forming a cycle and, each cycle is a wave. Therefore, this work will consider a complete contagion wave when there is a contamination cycle in a given region, that is when that region presents an increasing contagion curve, followed by a constant decline.

For each comparable country, the graph of new cases was elaborated, identifying the present cycles that can be observed in Figure 5.

**Figure 5.**
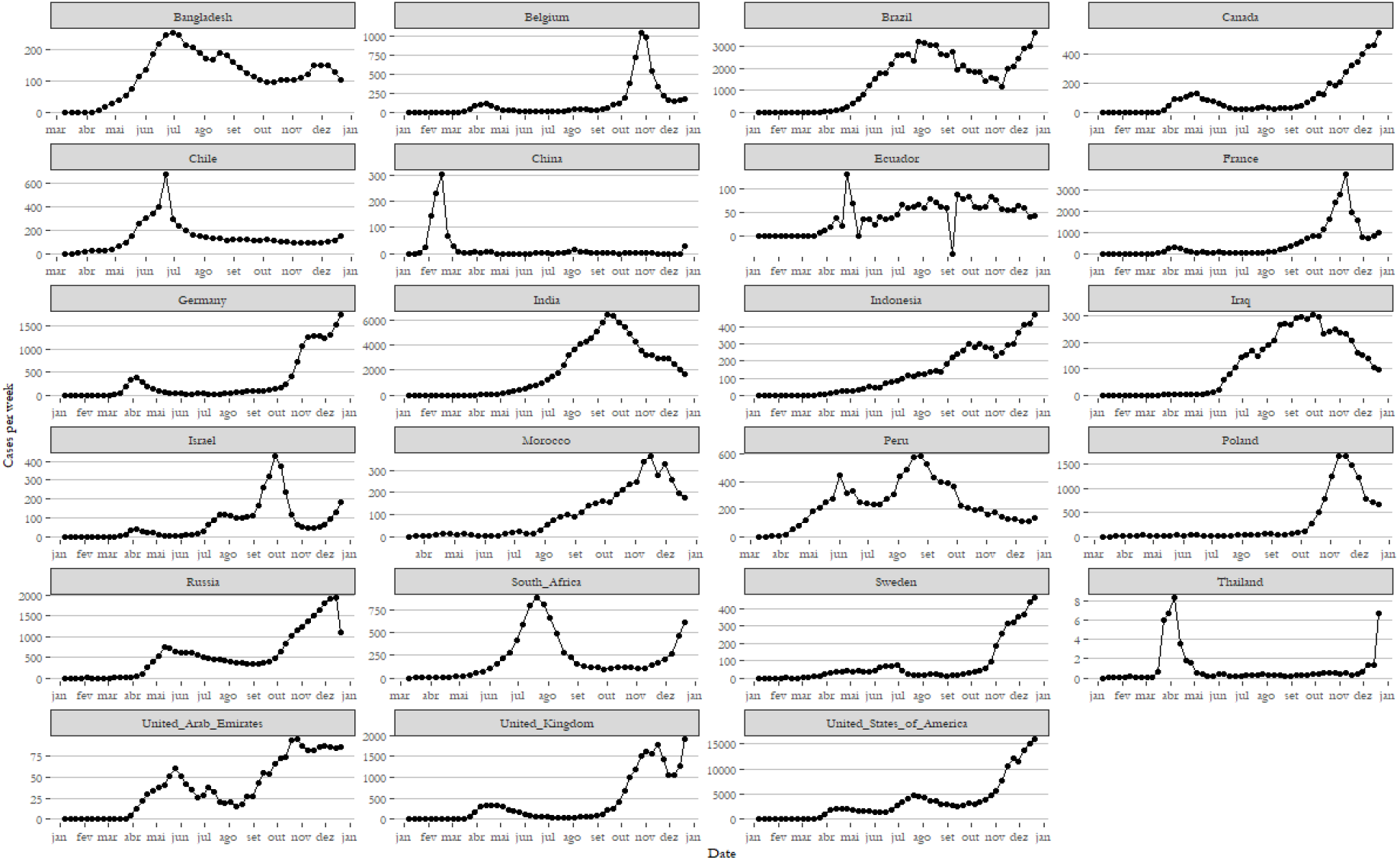
New cases (cases x 100). Source: author’s elaboration. Data: Our World in Data – Coronavirus (COVID-19) Cases, 2020.

Identifying contagion waves is important because, in some countries like Canada, United Arab Emirates, and the USA, the second wave is worse than the first one, exceeding the number of new cases of the first wave. To input, the right data in the model, identify the waves are necessary to establish two peaks, the first one for the first wave and the second one for the second wave.

Until mid-December, the peak days for comparable countries can be seen in Figure 6. With the peak days in possession, for the first and second waves, it is possible to start the MCM calculations using Equations 1 to 4.

**Figure 6.**
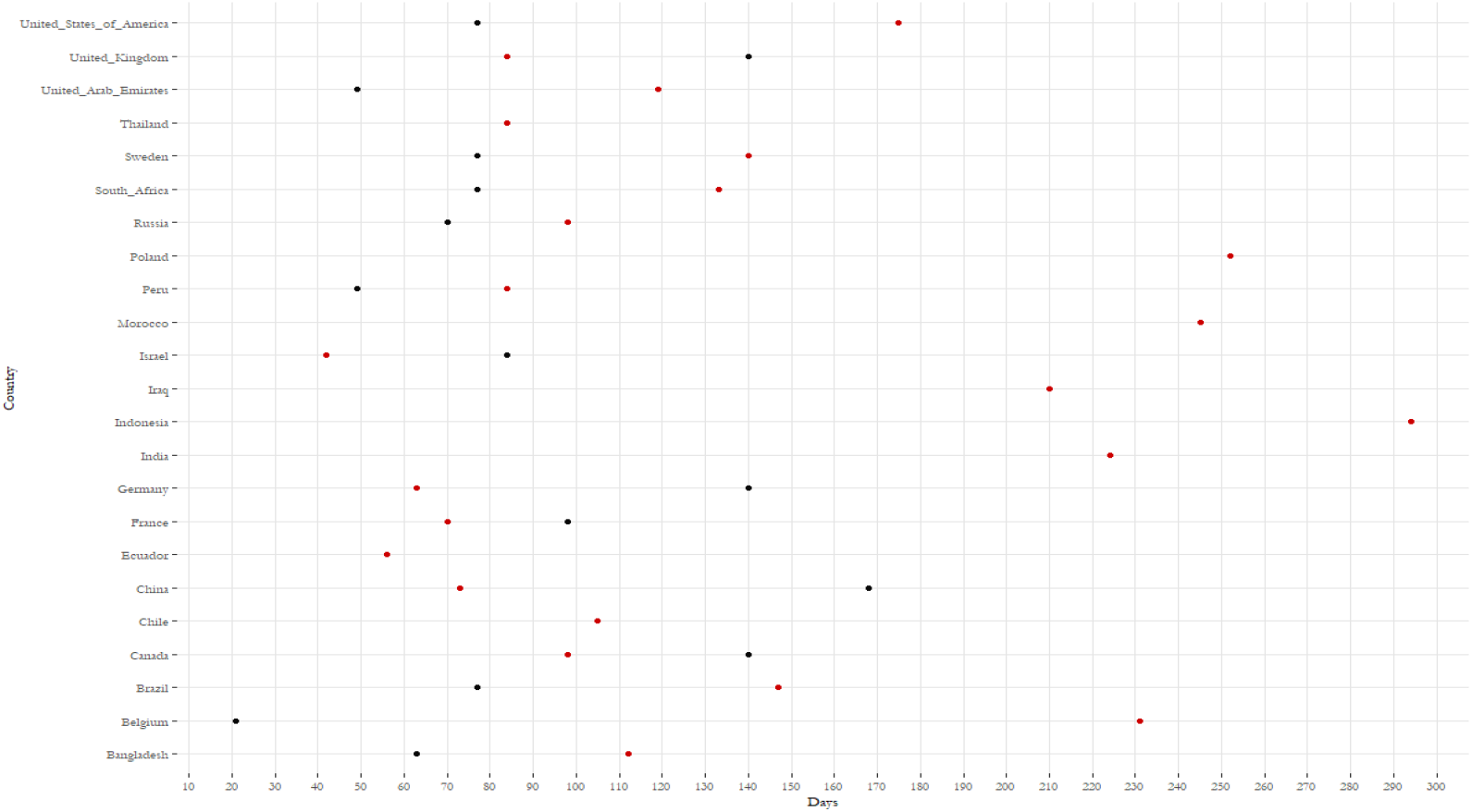
Days since first registered case for reach Covid-19 peak contamination. Note: red points are for the first wave and black points are for the second wave. Source: author’s elaboration.

However, this is a heterogeneous data sample in terms of climate and regional development. Several authors such as Antony and Rao (2007), Bettcher and Lee (2002), Alin and Marieta (2011), and Ghoncheh *et al*. (2016) indicate that the Human Development Index (HDI) interferes with public health, where the highest index reveals better quality health.

Still, regarding the heterogeneity of the countries, there is the climate factor. In most cases, climate interferes with illnesses like respiratory diseases. D’Amato *et al*. (2010, 2014), Lilleør and Van den Broeck (2011), Takizawa (2011), and Joshi *et al*. (2020) indicate that the climate has a directed impact on respiratory diseases and allergic respiratory disease. As climate impact factor was set to 1 for a similar climate and 0.75 for moderately similar and 0.5 for a different climate. This is an important driver to evaluate this type of disease and linked to Covid-19, which is a respiratory disease.

Knowing this fact, an outline of the countries was structured by the HDI and a climate impact factor. To do this, a cross-country regression was structured to equalize the countries by your angular coefficients. to structure this correlation, the precepts indicated in the studies of Levine and Zervos (1993) and Fernandez *et al*. (2001) whose main precepts are indicated by Equations 5 to 8.

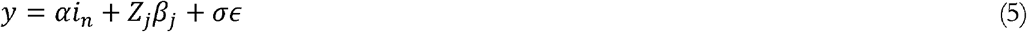

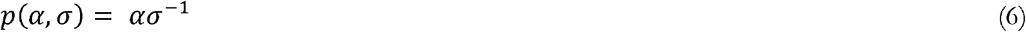

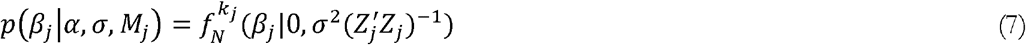

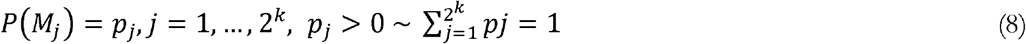

According to Levine and Zervos (1993) and Fernandez *et al*. (2001) cross regression to model the variables and coefficients follow the Bayesian statistical pattern and the model can be adapted to predict uncertainty, given by the posterior model probabilities which in this case is the MCM model. To equalize the comparable countries, the parameters *β* and *p* were used as HDI and climate, respectively, parameterizing the data imputed in the model.

To equalize the data between the selected countries, the climate factor impact and the HDI were used as shown in Table 1.

**Table 1.** Climate factor and HDI for the selected comparable countries. Source: Research data and United Nations Development Programme, 2015. Note: the climate impact factor was structured arbitrarily by the authors.

| **Comparable selected countries** | **Climate factor** | **HDI** |
| --- | --- | --- |
| Bangladesh | .75 | .570 |
| Belgium | .75 | .890 |
| Brazil | 1.0 | .755 |
| Canada | .50 | .915 |
| Chile | .75 | .832 |
| China | .75 | .755 |
| Ecuador | 1.0 | .732 |
| France | .50 | .888 |
| Germany | .50 | .916 |
| India | 1.0 | .609 |
| Indonesia | 1.0 | .684 |
| Iraq | .75 | .654 |
| Israel | .75 | .894 |
| Morocco | 1.0 | .628 |
| Peru | 1.0 | .734 |
| Poland | .50 | .843 |
| Russia | .50 | .798 |
| South Africa | .75 | .666 |
| Sweden | .50 | .907 |
| Thailand | .75 | .726 |
| United Arab Emirates | .75 | .835 |
| United Kingdom | .50 | .907 |
| United States of America | .75 | .915 |

After unleveraged the peak day for all selected counties the results are shown in Table 2.

**Table 2.** Climate factor and HDI for the selected comparable countries. Source: Research data.

| Countries | Peak 1<sup>st</sup><br>wave | Peak 2<sup>nd</sup><br>wave | Equalized<br>peak 1<sup>st</sup> wave | Equalized<br>peak 2<sup>nd</sup> wave |
| --- | --- | --- | --- | --- |
| Bangladesh | 112 | 63 | 73 | 41 |
| Belgium | 231 | 21 | 211 | 19 |
| Brazil | 147 | 70 | 110 | 52 |
| Canada | 98 | 133 | 93 | 127 |
| Chile | 105 | - | 91 | - |
| China | 73 | 28 | 59 | 22 |
| Ecuador | 56 | - | 40 | - |
| France | 70 | 98 | 65 | 92 |
| Germany | 63 | 133 | 60 | 127 |
| India | 224 | - | 136 | - |
| Indonesia | 287 | - | 196 | - |
| Iraq | 210 | - | 152 | - |
| Israel | 42 | 84 | 38 | 77 |
| Morocco | 245 | - | 153 | - |
| Peru | 84 | 49 | 61 | 35 |
| Poland | 252 | - | 231 | - |
| Russia | 98 | 70 | 87 | 62 |
| South Africa | 133 | 70 | 98 | 51 |
| Sweden | 140 | 70 | 133 | 66 |
| Thailand | 84 | - | 66 | - |
| United Arab Emirates | 119 | 49 | 103 | 42 |
| United Kingdom | 84 | 105 | 79 | 99 |
| United States of America | 175 | 70 | 163 | 65 |

Equalizing the data by the character of each country, it is intended to obtain raw data without leverage. This unleveraged data will serve to feed the MCM and BM model without the data being biased. After performing the MCM with unleveraged data, to apply the model in a given country, it is necessary to leverage the MCM result by the factors of the target country.

To performing the MCM method, a normality test needed to be executed. The normality test is indicated as knowing the behavior of the sample because the MCM is based on a standard normal distribution, therefore the contamination day data must follow this same distribution. Consequently, Siegel and Castellan Jr (1988) and Hair *et al*. (2007) indicate the Kolmogorov-Smirnov test or the Shapiro-Wilk test to investigate the normality of de dataset. Because of the large date, N>50, the Shapiro-Wilk test is recommended by Hair *et al*. (2005).

The test statistic can be represented using Equation 9, The test statistic can be represented through Equation 1, whose *F*_0_ is the accumulated theoretical distribution and *S*_*n*_ is the distribution observed in the equalized data.

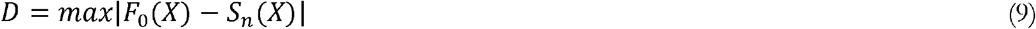

For the normality test, *H*_0_ is the null hypothesis: *H*_0_, distributions are significantly similar (same behavior); *H*_1_, distributions are not significantly similar (uneven behavior). The results for the Kolmogorov-Smirnov and Shapiro-Wilk test for each comparable country are present in Table 2.

With the normality tests performed, it was proved that the data of the number of days for the peak of contagion follows a normal distribution, accepting the hypothesis *H*_0_ of the Shapiro-Wilk test. Having proven normality, the MCM model of the normal distribution can be more accurately calibrated.

Observing the behavior of the contagion curve, Figure 5, is also a factor for an adequate calibration of the model, especially when input data in Equations 5 to 8.

An important part of the study is to cross-validate the results pointed on the MCM model. For this, Efron and Tibshirari (1985, 1997) indicate the bootstrap method that has the simple principle of Equation 10.

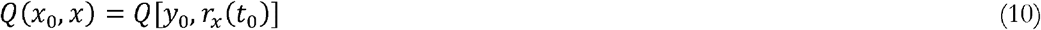

The bootstrap method used in this study is described in Efron and Tibshirari (1985, 1997) and will be used after obtaining the first frequency distribution of the MCM model, valid for first and second waves independently.

Combining all the calculation steps and the transformations in the database described in this section, the BMESPC model is presented.

### Covid-19 peak contamination in Brazil

Following the analytical approach, the BMESPC was performed to structure a distribution due to analyze the peak of Covid-19 contagion. The raw data was treated and the MCM and BM methods returned a probability and frequency distribution for peak days – first and second wave – shown in Figure 7.

**Figure 7.**
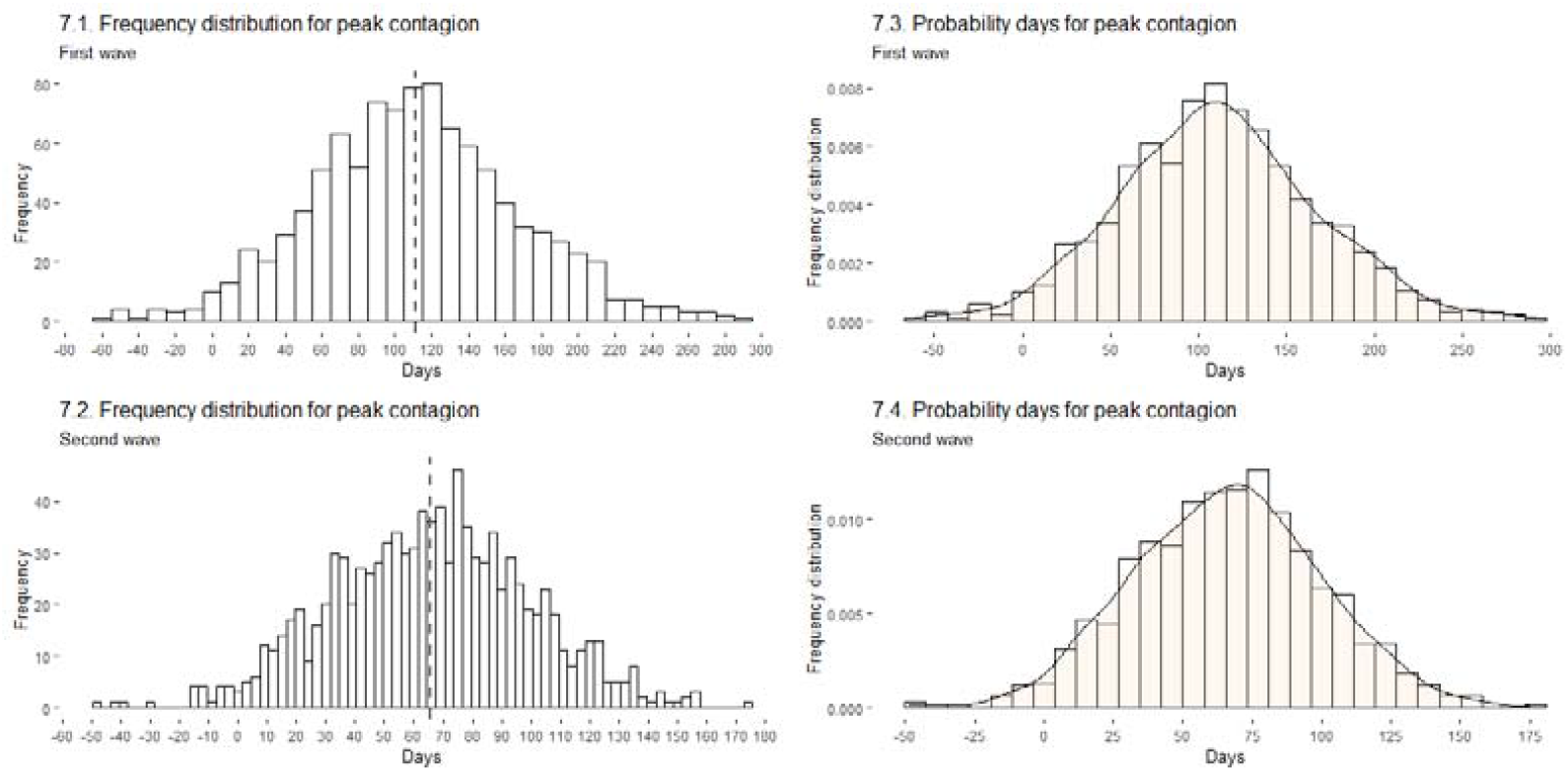
MCM and BM distribution of frequencies (in days). Source: Research data.

This probability distribution is the standard basis structured by the forecast model. To reflect reality, the base must be set according to the country of reference, in this case, Brazil. Subsequently, the average incubation period is added, in this study, the 14-day incubation period was used. However, Zhao *et al* (2020) estimated that the transmission may still occur during the incubation period, being the contagion by people who did not manifest any apparent symptoms.

Note that this study attempts to forecast the contagion date, which is approximately 14 days before the case is recorded due to the virus incubation period. Thus, the frequency distribution of Figure 7 indicates the period of contagion, pre-notification of new cases.

For this base data, the peak forecast for SARS-CoV-2 contagion for the first wave is on the 111^th^ day since the first case and may occur between the 73^rd^ and 145^th^ days. The forecast for the second wave of peak contagion is on the 65^th^ day after the end of the first wave which may occur between day 42^nd^ and 88^th^.

To find the distribution for Brazil it is necessary to leverage the data to the Brazilian standard, considering the same factors of HDI and climate as before. Thereby, the distribution for Brazil’s first wave of Covid1-9 contagion is indicated in Figure 8.

**Figure 8.**
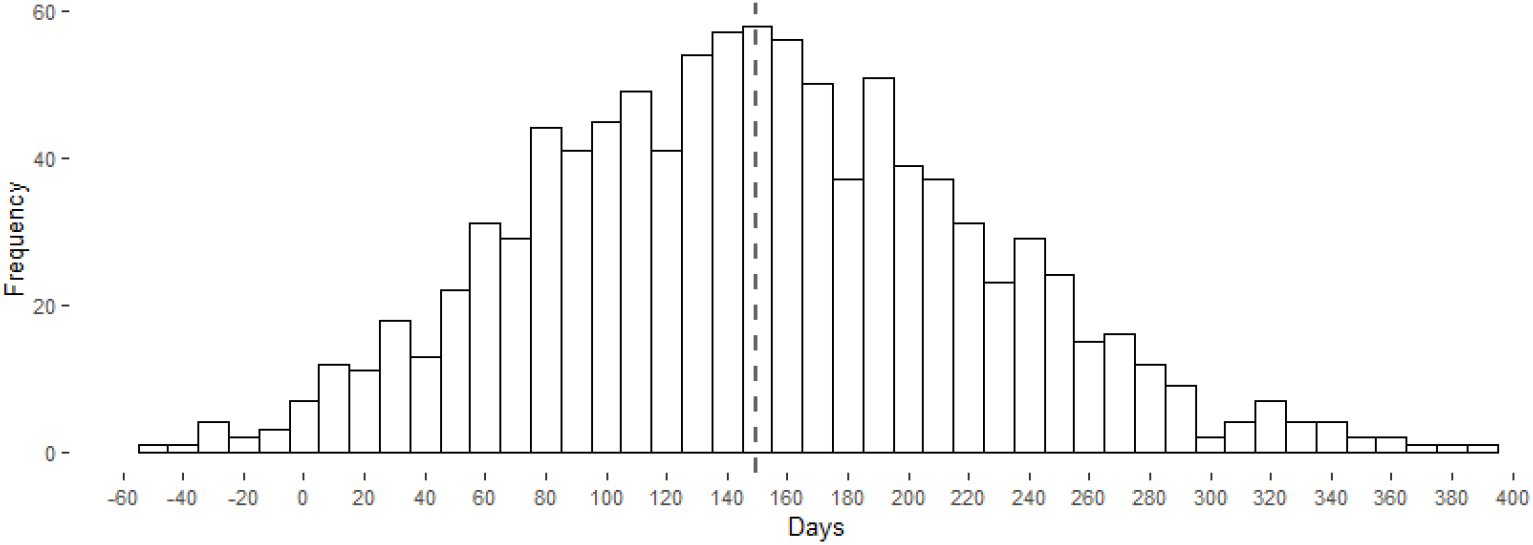
Brazilian’s BMESPC distribution of frequencies for first wave (in days). Source: Research data.

Contrary to previous predictions by Kuroiva *et al*. (2020a, 2020b, 2020c), this update shows that the peak of contagion in Brazil exceeds 100 days. Therefore, after a recalibration of the structured model and some statistical tests, the previous studies are revisited and corrected by this present study.

For the projected scenario, the first wave of SARS-CoV-2 contagion peak occurs between the 97^th^ and 197^th^ day after the first case with a forecast for the 149^th^ day. However, when observing the Brazilian data, the first wave peak occurred on July 23, that is, 148^th^ days of the first case, one day difference from the model.

By the minimum difference, it is possible to infer that the BMESPC is properly calibrated and running with the appropriate data.

Starting with the projection of the second wave in Brazil, Figure 9 indicates the distribution obtained by BMESPC.

**Figure 9.**
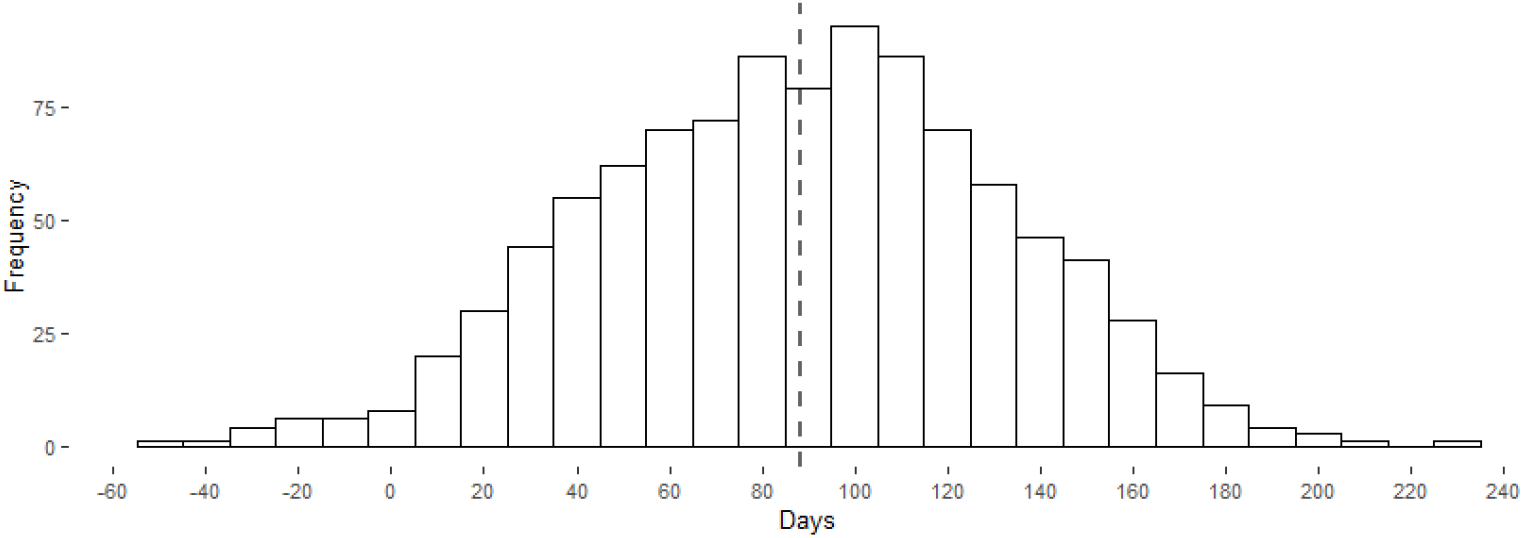
Brazilian’s BMESPC distribution of frequencies for a second wave (in days). Source: Research data.

The peak forecast for the second wave of contagion may occur between the 57^th^ and 118^th^ day after the end of the first wave. The estimation indicates the 88^th^ day as a peak of contagion which is given in a shorter time than the first wave. The decrease in the time that the peak is reached during the second wave is seen by other countries like the United Kingdom, Germany, and the USA.

With the forecast of the peak of the second wave for the 88th day since the end of the first wave (according to the model, the first wave in Brazil ended at the end of October, after the sharp drop in the number of cases as indicated by Figure 2), the peak will occur in mid-January.

Because Brazil has a vast territory, each one with different characteristics, the peak may vary from state to state. The distribution found in Figures 8 and 9 is valid for the country as a whole. the analysis by the state must have an adaptation since each one is in a different stage of the pandemic (for example different data from the first recorded infection).

At the state level, which one has your first case date and different characteristics that directly interfere with peak contamination. Following the same principles in BMESPC, the forecast for the state-level was carried out and the results are described in Table 4.

**Table 3.** Normality tests between comparable countries after equalization (95% confidence level). Source: Research data.

| Countries | Kolmogorov-Smirnova |  |  | Shapiro-Wilk |  |  |
| --- | --- | --- | --- | --- | --- | --- |
|  | Statistic | df | Sig. | Statistic | df | Sig. |
| Bangladesh | .167 | 127 | .258 | .866 | 127 | .233 |
| Belgium | .260 | 202 | .507 | .680 | 202 | .239 |
| Brazil | .259 | 202 | .007 | .740 | 202 | .161 |
| Canada | .186 | 202 | .646 | .834 | 202 | .627 |
| Chile | .250 | 137 | 1.977 | .509 | 137 | 1.741 |
| Ecuador | .331 | 197 | 1.740 | .405 | 197 | .414 |
| France | .252 | 202 | 1.676 | .703 | 202 | 1.097 |
| Germany | .283 | 202 | .024 | .667 | 202 | .106 |
| India | .266 | 201 | 1.422 | .687 | 201 | .410 |
| Indonesia | .204 | 195 | .441 | .812 | 195 | 1.317 |
| Iraq | .360 | 200 | 2.943 | .613 | 200 | .515 |
| Israel | .275 | 199 | .038 | .649 | 199 | .047 |
| Morocco | .131 | 132 | .786 | .841 | 132 | .130 |
| Peru | .119 | 134 | .776 | .911 | 134 | .230 |
| Poland | .103 | 136 | .123 | .954 | 136 | .173 |
| Russia | .261 | 202 | 1.774 | .797 | 202 | 1.724 |
| South Africa | .249 | 134 | .885 | .708 | 134 | .533 |
| Sweden | .213 | 202 | .051 | .775 | 202 | .249 |
| Thailand | .342 | 195 | .100 | .474 | 195 | .098 |
| United Arab Emirates | .229 | 196 | 2.174 | .858 | 196 | 1.507 |
| United Kingdom | .299 | 202 | .702 | .551 | 202 | 1.545 |
| United States of America | .211 | 202 | 1.224 | .857 | 202 | .825 |

**Table 4.**
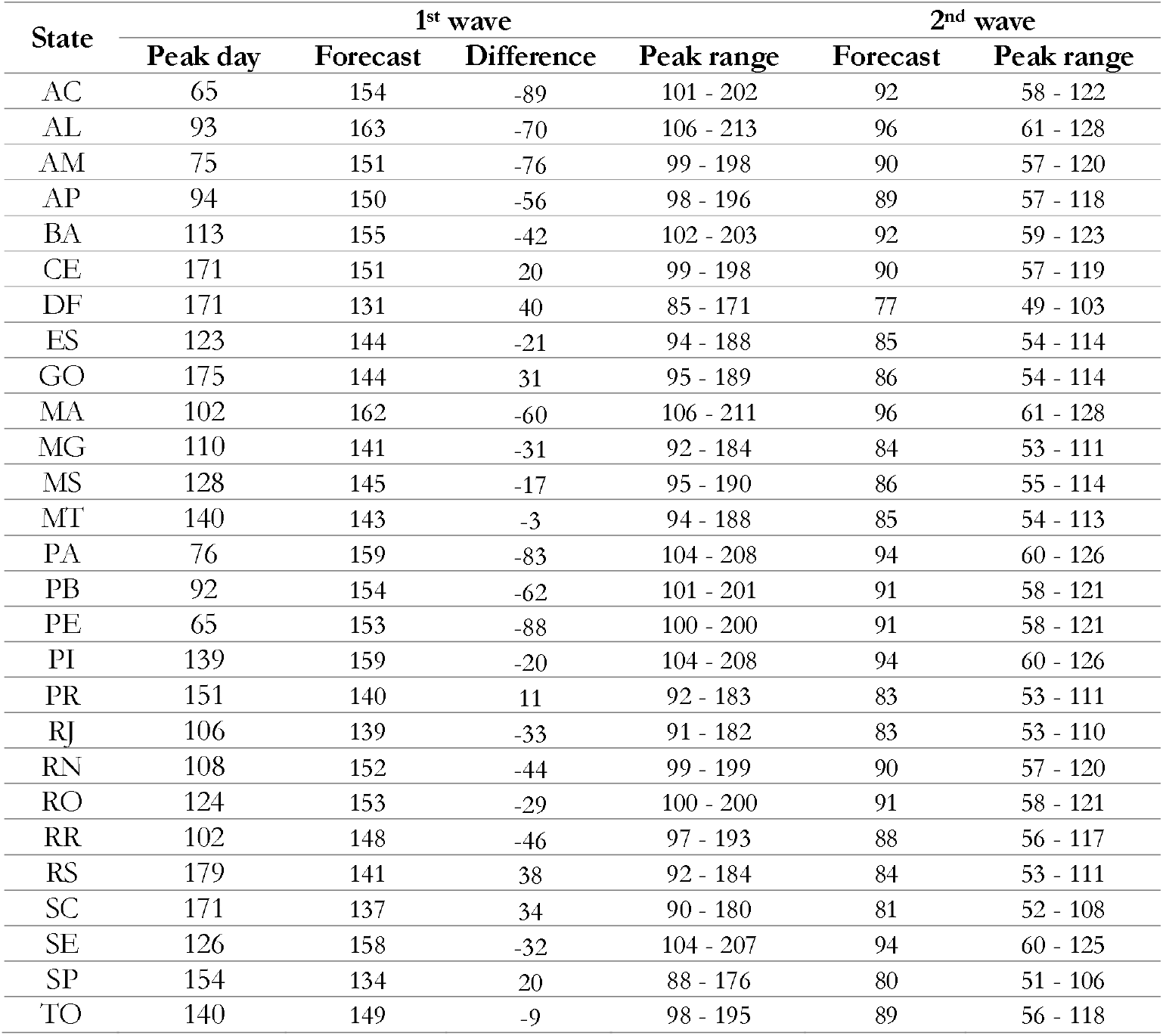
Days for peak contamination at the Brazilian state level. Source: Research data.

According to Table 4, in many states, the 1^st^ wave peak happened ahead of the BMESPC model forecast. In states like Acre (AC), Pará (PA), and Pernambuco (PE) the peak of contamination appeared much faster than expected. This is an indication that the virus spread rapidly in these states, at a more aggressive rate than in the others.

In contrast to the states that peaked earlier (for 1^st^ wave) than expected, there were states such as São Paulo (SP), Santa Catarina (SC), Paraná (PR), Goiás (GO) Federal District (DF), and Ceará (CE) that peaked beyond expected. Having this behavior indicated that there was a flattening of the contagion curve. For the other states that peaked earlier, this flatter did not happen.

It is worth mentioning that it is a probabilistic model that can be changed by exogenous variables, such as public policies and human behavior. These are some variables that contribute to the peak oscillation between one update to another. The probabilistic models demonstrate trends and not absolute truth.

Presenting a difference between what happened and what is expected is common when it comes to Bayesian statistics from which only data is analyzed and not other inferences such as public policies and human behavior. By just analyzing the data and making inferences, deviations in the values can take place, so a margin is stipulated (be expecting) the event to occur. So, in some cases, the peak is still within the predicted range.

Still concerning the difference between the actual dates and the predicted dates, other variables exogenous to the model influence to have this difference. Public policies, the rate of physical isolation, the socio-economic profile, and the demographic density are some examples. Among these characteristics, the rate of physical isolation, which derives from human behavior, is the one that most interferes, since in many Brazilian states there is no respect for the recommendation to remain at a social distance, contributing to the peak occurring before the forecast and faster.

Such factors are not present in the model that can explain the disparity between the real and the predicted. It is evident that human behavior, concerning social isolation, is different in the state of Acre from the state of São Paulo, for example. Just as the distance from the state of Pernambuco is different from the Chinese’s physical distance. For these reasons, there is a difference between the model. However, it is noted that, in general terms, the model is accurate when it comes to the country as a whole, demonstrated by the minimum difference of one day.

### Covid-19 peak contamination in Brazil

The objective of this work was to structure a model of forecasting the peak of contagion for the first and second waves in Brazil and that can be replicated for any region. For this, the Brazilian model estimation for the SARS-CoV-2 peak contagion model – BMESPC –was structured using Bayesian statistics and the Monte Carlo method for predicting events in uncertainty.

With the collected data from Our World in Data and the Brazilian Healthy Ministry sources it was possible to structure and calibrate the BMESPC model. The model proved to have limited accuracy due to the large discrepancy in raw data (for Brazil the data referring to Covid-19 are incipient) and the socio-economic-political-cultural difference between states. However, in the case of Brazil, the accuracy of the model proved to be correct, with one day difference between the forecast and real, in the case of the first wave. It is noteworthy that, even with a difference in the projection and the real, the occurrence interval remains correct in most cases.

Therefore, the BMESPC model has its usefulness in predicting the peak of contagion, being able to be replicated for several regions as long as the proper structure and calibration is followed.

## Data Availability

All data was collected from thrid part websites.

https://www.ecdc.europa.eu/en/publications-data/data-national-14-day-notification-rate-covid-19

https://covid.saude.gov.br/

## Notes

### Competing Interest Statement

The authors have declared no competing interest.

### Funding Statement

No funding for this paper.

### Author Declarations

No approval needed for this paper.

### Summary of Updates

Update for Brazilian States and title.

